# Determinants of measles second dose vaccination dropout among children 24 – 35 months of age in Addis Ababa, Ethiopia. 2025: A Community based Unmatched Case-Control study

**DOI:** 10.64898/2026.02.10.26346050

**Authors:** Belete Getachew, Zewdie Aderaw, Demeke Taye, Simegnew Handebo

## Abstract

**Background:** Measles is a highly contagious infectious disease and a leading cause of childhood morbidity and mortality worldwide. In developing country like Ethiopia, effective immunization is a proven strategy for reducing measles related illness and deaths. However, measles second dose vaccination drop out has become a major public health concern. In a densely populated city such as Addis Ababa drop rate tends to be higher than the minimum acceptable threshold, leading to increased number of cases and recurrent outbreaks. Despite of this limited evidence exists on the determinants of second dose drop out and the problem is not well investigated, as a result this study will try to identify determinants of measles second dose vaccination dropout among children 24 – 35 months of age.

**Objectives:** To identify determinants of measles second dose vaccination dropout among children 24 – 35 months of age Addis Ababa, Ethiopia in 2025.

**Method:** Community based unmatched case control study was conducted in Addis Ababa from September 1/2024 to October /2025 with a total of 636 participants, consisting of 212 cases and 424 controls. Data were collected using structured Quesionariie and entered into EpiData 3.1 then StataSE 18 was used for detailed analysis including Descriptive statistics. Model fitness was checked using Hosmer-Lemeshow and multicollinearity were assessed using variance inflation factor. Furthermore, Bivariable and multivariable logistic regression analyses was employed and Adjusted odds ratio with 95% confidence intervals was used to identify significant variables.

**Results:** A total of 620 mothers/caregivers participants respond to the study, comprising 206 (97%) cases and 414(97.6%) controls, yielding a total response rate of 97.4%. In this study, waiting time longer than 30 minutes (AOR= 3.34, 95%CI: 1.86-5.9), Lack of counseling (AOR = 2.63, 95% CI: 1.60–4.30), Lack of reminders (AOR = 2.86, 95% CI: 1.89–4.30), Previous adverse event following immunization (AOR = 2.00, 95% CI: 1.39–3.00), postnatal care visit (AOR = 0.58, 95% CI: 0.40–0.85) and family size of greater than 3 (AOR = 1.96, 95% CI: 1.29–2.98) were significantly associated with measles second dose dropout.

**Conclusion and recommendation:** In study shows measles second dose dropout is found to be associated with long waiting time, lack of counseling, lack of reminder, history of adverse event following immunization and postnatal visit. Which suggests Strengthening Immunization Counseling, reducing waiting time, establishing effective reminding system, integrating Immunization with postnatal services and promptly addressing concerns about adverse event following immunization can help reduce measles second dose dropout.

## Introduction

Childhood immunization is one of the most effective public health interventions to reduce morbidity and mortality from vaccine preventable diseases (1). Among those disease measles remains a critical focus due to its highly contagious nature and severe complications (2). According to World health organization (WHO), the number of measles cases globally rose from over 170,000 in 2022 to over 320,000 in 2023, making it one of the primary causes of child morbidity and mortality worldwide. Additionally, there have been almost 100,000 measles cases in the first few months of 2024. Sub-Saharan Africa and Southeast Asia are home to the majority of those cases(3,4).

The global effort toward measles control and elimination began in the 1960s with the development and widespread use of the measles vaccine. Over decades, numerous strategies have been implemented to increase vaccination coverage, including introduction of 2^nd^ dose, immunization awareness campaigns, and strengthening Expanded Program on Immunization (EPI) (3,5–8).

The measles containing vaccine (MCV) is one of the most effective and cost-efficient tools to prevent measles. The World Health Organization (WHO) recommends two doses of MCV as part of routine immunization. The routine administration of a second dose of measles containing vaccine (MCV2), usually in the second year of life (2YL) between 15 and 18 months of age, reduces susceptibility in up to 15% of individuals who do not seroconvert after the first dose of MCV at 9 months of age (9).

Global coverage of MCV1 and MCV2 is approximately 81% and 71%, respectively with a dropout of 12%, as of 2022. In the WHO African Region, in 2022, 69% MCV1 and 45% MCV2 coverage was estimated with a dropout of approximately 35%. From all countries where MCV2 is included in routine immunization schedules for over three years, 23 countries including Ethiopia experienced a dropout of over 10% which is above the WHO tolerable level (10,11). Such a large dropout rate suggests problems in completing the measles vaccine series among children. The overall measles vaccination dropout rate in Ethiopia was 33.3% (95% CI: 31.6, 34.9). The urban and rural measles vaccination dropout rates in Ethiopia were found to be 34% and 66%, respectively(12).

Ethiopia has actively engaged to improve MCV2 coverage such as incorporating MCV2 into its routine immunization program in 2019, conducting mass immunization campaigns, Community engagement, leveraging religious and traditional leaders to reduce vaccine hesitancy and Integration with maternal and child health services. Despite these activities COVID-19 pandemic, unsustainability of immunization campaigns, misinformation and cultural barriers, reliance on donors lags its implementation and leaves children’s drop out of MCV2. Additional challenges, such as vaccine stockouts, cold chain disruptions, and insufficiently trained healthcare workers, exacerbate the problem. This challenges not only leave children vulnerable to outbreak but also placing a significant barrier to Ethiopia achieving the WHO’s target of at least 95% coverage for both MCV doses by 2025 (9,13).

In Addis Ababa, where population density is high, low MCV2 coverage could easily lead to outbreaks, placing additional strain on the public health system. Moreover, incomplete vaccination weakens herd immunity, which is essential in protecting individuals who are too young or medically unable to receive the vaccine. Thus, high MCV2 dropout rates in Addis Ababa not only threaten the health of individual children but also increase the risk of outbreaks within the community. Furthermore, it also undermines the cities potential to reach herd immunity, making each community more susceptible to outbreaks.

A review of existing literature indicates that MCV2 dropout is influenced by factors such as limited parental knowledge, access to healthcare, and socioeconomic disparities(14), Since Addis Ababa has unique features like high internal immigration different socio economic and cultural perspective and overwhelming health system affected by COVID-19, the specific determinants in urban settings remain inadequately understood (15).

Addressing these factors and closing the MCV2 gap is essential to achieving the targets set by Ethiopia’s national immunization program and the broader goals of the WHO’s Global Immunization agenda2030, which aims for high and equitable immunization coverage globally (8,16,17). As a result, this finding will help inform public health strategies to reduce dropout rates, enhance MCV2 coverage, and ultimately contribute to measles prevention efforts in Addis Ababa.

## Methods and materials

### Study setting/area and period

Addis Ababa is in the heartland of the country in an area of 540 square kilometers. It is situated between 9 degrees’ latitude and 38 degrees’ east longitude in a plateau that stretches at the range of 2200-2800 meters of altitude above sea level. The Climate varies from seasons of summer, about 9 months, to cool months of rainfall, about 3 months, with an overall average, the maximum and minimum temperature of 22.9 and 10.8 degrees centigrade, respectively and total population 4,104,864 of them 52% females and 48% are male. The city has 11 sub city, 118 woreda, 96 health centers and 14 public hospitals There are over 1047 health facilities in Addis Ababa of which 937 of them are private facilities(18). A study was conducted from September 2024 to October 2025.

### Study design

A community-based unmatched case control study design was used to investigate the determinants of measles second dose vaccination dropout among children 24 – 35 months of age Addis Ababa, Ethiopia 2025.

### Source population

Cases: All children aged 24-35 months residing in Addis Ababa who received the first dose of measles-containing vaccine (MCV1) but did not receive the second dose (MCV2).

Controls: All children aged 24-35 months residing in Addis Ababa who received both MCV1 and MCV2.

### Study population

Cases: All children aged 24-35 months living in selected woreda and identified as received MCV1 but not MCV2.

Controls: All children aged 24-35 months living in selected woreda who completed both MCV1 and MCV2

### Eligibility criteria

Inclusion criteria Cases

Children aged 24-35 months.

Children parent residing in Addis Ababa for at least six months. Received MCV1 but did not receive MCV2.

Controls

Children aged 24-35 months.

Residing in Addis Ababa for less than six months. Received both MCV1 and MCV2.

Exclusion criteria

Cases

Children aged below 24 and above 35 months.

Children parents not Residing in Addis Ababa for less than six months. Children whose vaccination records are missing or incomplete.

Controls

Children aged below 24 and above 35 months.

Children parents not Residing in Addis Ababa for less than six months Children whose vaccination records are missing or incomplete.

Children receive both MCV1 and MCV2 through campaign

### Sample size determination

The sample size was calculated by using Epi info version 7.2.6.0 Stat Calc. from study conducted in East Bale Zone, Ethiopia. determinant factors for measles second dose drop out were Mothers who were unable to read and write, did not receive counseling, spent ≥30 min to reach health facilities, and did not attend postnatal care, mothers who had poor knowledge of second-dose measles waited more than an hour for measles vaccination at health facilities were significantly associated with second-dose measles vaccination drop out (19).

Therefore, the required sample size was 386, After accounting for a 10% non-response rate and a design effect of 1.5 the final estimated sample size became 636, consisting of 212 cases and 424 controls.

### Sampling technique

Firstly, Multistage sampling technique was used to select the study subjects Firstly Four sub cities was selected using simple random sampling technique, then from each sub city Yeka (4), Gulele (3), Akaki kality (3), and Lemi kura (3) corresponding woreda was selected.

Secondly, the study children were selected by simple random sampling technique (lottery method) using a sampling frame prepared from a list of cases and controls obtained with full address from EPI registration data of the catchment health facility.

### Study Variable

Dependent variable Measles second dose drop-out

#### Independent variables

The independent variables that associated to measles second dose immunization drop-out include: Socio-Demographic Characteristics of Mother/Caregiver: - Age of the mother/caregiver, Sex of the caregiver, Family size, educational status of the caregiver, Occupation, Marital status, Knowledge of Mother/Caregiver, Knowledge, Awareness of MCV2 importance, Fear of side effects

Child-Related Factors: - Sex of the child, Child illness (recent or chronic), Age of the child at vaccination, Fear of side effects related to the child and Number of children vaccinated per session (perceived impact on the child

Health System Level Factors: - Availability, accessibility, and quality of immunization services, Vaccinator absence, Vaccine availability (stockouts), Long waiting times, Distance to health facility

#### Operational definitions

Measles second dose dropout: - children who took routine MCV1 at 9 months but not took routine MCV2 at 24-35 months (19).

Knowledge: mothers who answered above median on knowledge questions was referred to as having good knowledge whereas answered the below median considered to have poor knowledge

Distance to nearest health facility: The distance between health service user’s/ of community residence area and immunization provision site takes >30 minutes in foot walking classified far and 30 minutes and below is classified as acceptable (not far) distance (20).

Waiting time for immunizing child: in this study refers mother who come health facility to vaccinate her child against after arrival waits >30 minutes is long waiting time and ≤ 30minutes is acceptable waiting time (21).

Adverse Event following Immunization (AEFI): Any untoward medical occurrence which follows immunization and does not necessarily have a causal relationship with the use of the vaccine(3).

#### Data collection tool, methods and procedures

WHO immunization survey reference manual, national & regional checklist and different literatures were adapted by addressing important variables including socio-demographic variables, child characteristics, accessibility of immunization service and maternal health service utilization related Factor(7,19,22–26). The questionnaire is prepared in English and then translated to Amharic.

#### Data quality control

To assure data quality Questionnaire was prepared in English by reviewing different literatures and translated in to Amharic (local language) and back to English by using two different language experts for consistency. Data collectors were trained. The questionnaire was pre tested on 5% of the sample size out of study area on population with similar characteristics in bole and Kirkos sub city two woredas.

The supervisors and principal investigator will closely follow the day-to-day data collection process to ensure completeness and consistency of the collected questionnaires on a daily basis. The data was checked manually, Coded and then entered into EpiData software. The data collectors were informed and interviewed Mothers or caretakers who had eligible children. They were submitting collected data regularly to supervisors to principal investigators. During data collection, mothers/caregivers who were not present to respond were revisited again in the next day.

#### Data processing and analysis

Prior to analysis, the whole data was cleaned and checked for completeness then the data was entered into Epi data version 3.1 and was exported to the Stata version 18 for further analysis. Data exploration was conducted to examine different characteristics of the data. By using Descriptive statistics, the frequency distribution of both dependent and independent variables was done. Hosmer-Lemeshow test was used to check goodness of fit of the model and Multicollinearity among independently associated variables was checked by Multicollinearity diagnostic test VIF.

Model fitness was assessed using the Hosmer–Lemeshow test, which indicated good fit (p = 0.425). Multicollinearity among the independent variables was tested using the Variance Inflation Factor (VIF). VIF value for all variables was within 1.04 and 3.33 and the mean was approximately 1.73. Since all the VIF values were well below the commonly applied threshold value of 5, this indicates absence of multicollinearity among the explanatory variables.

Binary logistic regression analysis was used primarily to check crude association of independent variables. Variables having an association with the outcome variable a p-value of <0.25 was candidate for multivariable logistic regression analysis. Adjusted odds ratio with corresponding 95% confidence intervals was used to show association between independent variables and a dependent variable. Those independent variables with P-value < 0.05 were considered statistically significant factors with outcome variable (measles dropout). Finally, results were presented in summary statics, text, graph and tables.

## Results

### Sociodemographic characteristics

In this study a total of 620 mothers/caregivers participated, comprising 206 (97%) cases and 414(97.6%) controls, yielding a total response rate of 97.4%.

According to the finding presented in Table 1, the mean age of mother/care giver is 31.37 years (SD ± 4.98) for cases 30.7 years (SD ± 4.94) and 31.7 years (SD ± 4.98) for controls. The majority of mother or care giver is on the age group between 25 and 34. On the other hand, the median age of the child is 29 months with interquartile range of 5. For cases the median age is 30 with 5 interquartile range, for control the median age is 29 with 5 interquartile range.

**Table 1:** Socio demographic characteristics of participants for determinants of measles MCV2 vaccination dropout among children aged 24-35 months of age, Addis Ababa Ethiopia 2025.

| Variables | Category | Case (n=206) | Control (n=414) | Total (n=620) |
| --- | --- | --- | --- | --- |

|  |  | Freq. | (%) | Freq. | (%) | Freq. | (%) |
| --- | --- | --- | --- | --- | --- | --- | --- |
| Mother/care giver Age (n=620) | 15-24 | 22 | 10.7% | 33 | 8.0% | 55 | 8.9% |
|  | 25-34 | 135 | 65.5% | 270 | 65.2% | 405 | 65.3% |
|  | 35 and above | 49 | 23.8% | 111 | 26.8% | 160 | 25.8% |
| Child sex (n=620) | Male | 110 | 53.4% | 194 | 46.9% | 304 | 49.0% |
|  | Female | 96 | 46.6% | 220 | 53.1% | 316 | 51.0% |
| Child age (month) (n=620) | 24-27 | 48 | 23.3% | 99 | 23.9% | 147 | 23.7% |
|  | 28-31 | 88 | 42.7% | 175 | 42.3% | 263 | 42.4% |
|  | 32-35 | 70 | 34.0% | 140 | 33.8% | 210 | 33.9% |
| Birth order (n=620) | 1st | 30 | 14.6% | 70 | 16.9% | 100 | 16.1% |
|  | 2nd | 45 | 21.8% | 81 | 19.6% | 126 | 20.3% |
|  | 3<sup>rd</sup> & above | 131 | 63.6% | 263 | 63.5% | 394 | 63.5% |
| Family size (n=620) | <=3 | 138 | 67.0% | 327 | 79.0% | 465 | 75.0% |
|  | >3 | 68 | 33.0% | 87 | 21.0% | 155 | 25.0% |
| Mother/care giver educational status (n=620) | Unable to read & write | 14 | 6.8% | 30 | 7.2% | 44 | 7.1% |
|  | Only read and write | 27 | 13.1% | 60 | 14.5% | 87 | 14.0% |
|  | Primary | 106 | 51.5% | 158 | 38.2% | 264 | 42.6% |
|  | Secondary & above | 59 | 28.6% | 166 | 40.1% | 225 | 36.3% |
| Marital status (n=620) | Single | 33 | 16.0% | 53 | 12.8% | 86 | 13.9% |
|  | Married | 100 | 48.5% | 202 | 48.8% | 302 | 48.7% |
|  | Separated/Divorced | 44 | 21.4% | 96 | 23.2% | 140 | 22.6% |
|  | Widowed | 29 | 14.1% | 63 | 15.2% | 92 | 14.8% |
| Occupational status of the mother (n=620) | Gov't employee | 27 | 13.1% | 48 | 11.6% | 75 | 12.1% |
|  | Private | 31 | 15.0% | 67 | 16.2% | 98 | 15.8% |
|  | House wife | 70 | 34.0% | 142 | 34.3% | 212 | 34.2% |
|  | Merchant | 33 | 16.0% | 63 | 15.2% | 96 | 15.5% |
|  | Student | 18 | 8.7% | 26 | 6.3% | 44 | 7.1% |
|  | Other | 27 | 13.1% | 68 | 16.4% | 95 | 15.3% |
| Mother/care giver knowledge(n=620) | Good | 97 | 47.1% | 218 | 52.7% | 315 | 50.8% |
|  | Poor | 109 | 52.9% | 196 | 47.3% | 305 | 49.2% |

In terms of educational status, 51.5% of cases and 38.2% of controls had completed primary education, while 28.6 % of cases and 40.1% of controls had attained secondary education or higher. The Occupation status finding revealed, housewives constituted 34% of cases and 34.3% of controls, followed by Merchant which constitutes 16% for cases and 15.2 % among controls.

#### Health care system related factors

The majority of respondents reported that their nearest health facility was a health center, accounting for 13.2% of cases and 33.5% of controls, followed by private health facilities.

About 53.9% of controls indicated that their nearest health facility was within a 30-minute walking distance. Conversely, comparable proportion of cases (12.6%) reported walking for more than 30 minutes to seek care. reach a facility. Furthermore, waiting time of > 30 minutes were more common among cases (91.3%) than Controls (79.2%)

A total of 372 (60%) of respondents indicates they have missed the vaccination schedule of which 145(38.9%) reported due to lack of reminder this is more common among cases (57.2%) than controls.

#### Child related factors

Among the respondents, 19.2% reported that their child was sick on vaccination day of which only 42% get advice from HCW. and 12.9% of controls indicated that their child was not sick at the time of vaccination. Among those whose child was sick on the vaccination day, 33.3% of cases and 46.3% of controls sought advice from health care workers.

Regarding attitudes toward immunization, 49% of cases and 43.5% controls reported fearing side effects, while a slightly higher proportion (51% of cases and 56.5% of controls) indicated no fear of side effects.

Adverse event following immunization from previous vaccination were indicated by 178(28.7%) of respondents which is more prevalent among cases (33.5%) than controls (26.3%). Among those who experienced AEFI, about 71% cases and 56.9% of controls said that this experience made them hesitant toward vaccination. Mother / care giver -related factors

Among the respondents, Regarding the place of delivery, more than half of the controls 51.6% and about one fourth (24.2%) delivered their children at a health center, while hospital accounted for 7.6% of cases and 12.9% of controls. Post natal visit was notably higher among controls (46.1%) compared to cases (18.7%), whereas lack of visit was more common among cases (14.5%) than controls (20.6%). Interms of perception towards the measles vaccine MCV2, lack of faith in the vaccine was reported by 9.7% of cases and 15.0% of controls, while the majority expressed confidence in MCV2 (23.5% of cases and 51.8% of controls)

Family support for vaccination was reported by 2.6% of cases and 4% of Controls, while the majority indicated a lack of family support (30.6% of cases and 62.7% of controls). Similarly concerns about misinformation regarding vaccination were minimal, with only 2.6% of cases and 4.2% of controls expressing the concern. Furthermore Table 2 shows work load as a barrier to vaccination by 92.2% of cases and 77.8% of controls, while 7.8% of cases and 22.2% of controls stated that their workload did not interfere with child vaccination.

**Table 2:** Mother / Care giver related determinants of measles MCV2 vaccination dropout among children aged 24-35 months of age, Addis Ababa Ethiopia 2025.

| Variables | Category | Case (n=206) |  | Control (n=414) |  | Total (n=620) |  |
| --- | --- | --- | --- | --- | --- | --- | --- |
|  |  | Freq. | (%) | Freq. | (%) | Freq. | (%) |
| Place of Delivery<br>(n=620) | Hospital | 47 | 22.8% | 80 | 19.3% | 127 | 20.5% |
|  | Health center | 150 | 72.8% | 320 | 77.3% | 470 | 75.8% |
|  | Private HF | 9 | 4.4% | 14 | 3.4% | 23 | 3.7% |
| Attend PNC (n=620) | Yes | 116 | 56.3% | 286 | 69.1% | 402 | 64.8% |
|  | No | 90 | 43.7% | 128 | 30.9% | 218 | 35.2% |
| Lack of faith on MCV2<br>vaccination (n=620) | Yes | 60 | 29.1% | 93 | 22.5% | 153 | 24.7% |
|  | No | 146 | 70.9% | 321 | 77.5% | 467 | 75.3% |
| Family member Support<br>(n=620) | Yes | 16 | 7.8% | 25 | 6.0% | 41 | 6.6% |
|  | No | 190 | 92.2% | 389 | 94.0% | 579 | 93.4% |
| Concern of<br>misinformation (n=620) | Yes | 16 | 7.8% | 26 | 6.3% | 42 | 6.8% |
|  | No | 190 | 92.2% | 388 | 93.7% | 578 | 93.2% |
| Work load affect<br>vaccinating (n=620) | Yes | 190 | 92.2% | 322 | 77.8% | 512 | 82.6% |
|  | No | 16 | 7.8% | 92 | 22.2% | 108 | 17.4% |

**Table 3:**
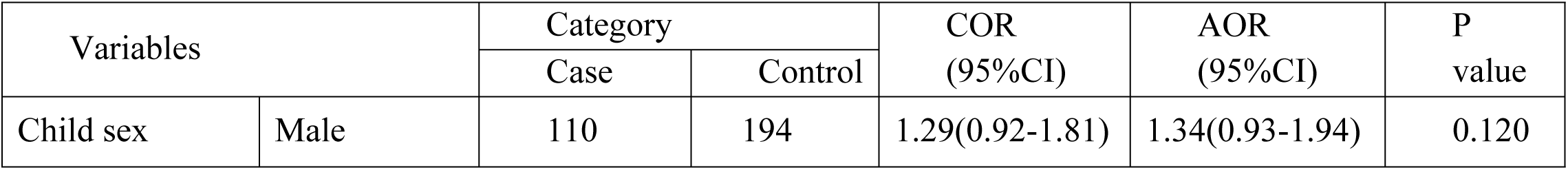
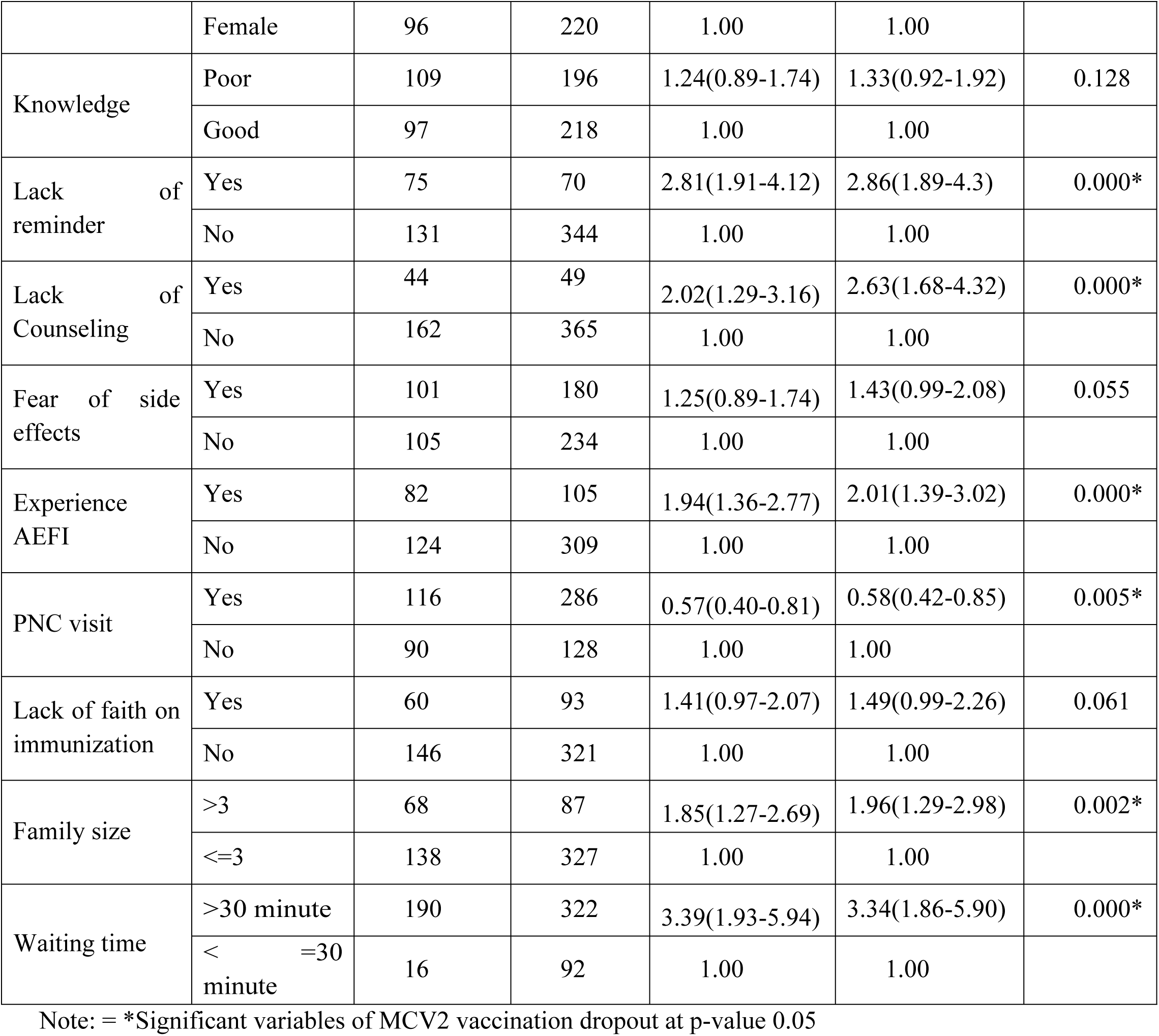
Bivariable and multivariable analysis of measles MCV2 vaccination dropout among children aged 24-35 months of age, Addis Ababa Ethiopia 2025.

| Variables |  | Category |  | COR<br>(95%CI) | AOR<br>(95%CI) | P<br>value |
| --- | --- | --- | --- | --- | --- | --- |
|  |  | Case | Control |  |  |  |
| Child sex | Male | 110 | 194 | 1.29(0.92-1.81) | 1.34(0.93-1.94) | 0.120 |
|  | Female | 96 | 220 | 1.00 | 1.00 |  |
| Knowledge | Poor | 109 | 196 | 1.24(0.89-1.74) | 1.33(0.92-1.92) | 0.128 |
|  | Good | 97 | 218 | 1.00 | 1.00 |  |
| Lack of reminder | Yes | 75 | 70 | 2.81(1.91-4.12) | 2.86(1.89-4.3) | 0.000* |
|  | No | 131 | 344 | 1.00 | 1.00 |  |
| Lack of Counseling | Yes | 44 | 49 | 2.02(1.29-3.16) | 2.63(1.68-4.32) | 0.000* |
|  | No | 162 | 365 | 1.00 | 1.00 |  |
| Fear of side effects | Yes | 101 | 180 | 1.25(0.89-1.74) | 1.43(0.99-2.08) | 0.055 |
|  | No | 105 | 234 | 1.00 | 1.00 |  |
| Experience AEFI | Yes | 82 | 105 | 1.94(1.36-2.77) | 2.01(1.39-3.02) | 0.000* |
|  | No | 124 | 309 | 1.00 | 1.00 |  |
| PNC visit | Yes | 116 | 286 | 0.57(0.40-0.81) | 0.58(0.42-0.85) | 0.005* |
|  | No | 90 | 128 | 1.00 | 1.00 |  |
| Lack of faith on immunization | Yes | 60 | 93 | 1.41(0.97-2.07) | 1.49(0.99-2.26) | 0.061 |
|  | No | 146 | 321 | 1.00 | 1.00 |  |
| Family size | >3 | 68 | 87 | 1.85(1.27-2.69) | 1.96(1.29-2.98) | 0.002* |
|  | <=3 | 138 | 327 | 1.00 | 1.00 |  |
| Waiting time | >30 minute | 190 | 322 | 3.39(1.93-5.94) | 3.34(1.86-5.90) | 0.000* |
|  | <=30 minute | 16 | 92 | 1.00 | 1.00 |  |

Note: = \*Significant variables of MCV2 vaccination dropout at p-value 0.05

#### Determinants of measles second dose vaccination dropout

Among the variables entered in Bivariable logistic regression, Child sex, Knowledge, Lack of reminder, Poor Counseling, Fear of side effects, experience AEFI, PNC visit, Lack of faith in vaccination, family size and waiting time were found to be a candidate for Multivariable logistics regression analysis with p value <0.25. from the above variables five of them were found to be statically significant with MCV2 Vaccination dropout shown in Table Mother/caregiver who didn’t receive reminder of vaccination were 2.86 time more likely to have the children drop out of MCV2 as compared to those who received reminders (AOR=2.86, 95%CI: 1.89-4.30). Similarly, poor Counseling During Immunization Visits was associated with more than 2.63 more likely to be drop out from MCV2 than those who weren’t received Counseling. (AOR=2.63,95% CI: 1.68-4.32).

Children who experienced an adverse event following immunization (AEFI) after the first dose had two times higher odds of dropout compared to those without such experience (AOR = 2.00, 95% CI: 1.39–3.00). In contrast, children of mothers who had attended postnatal care (PNC) visits were significantly 42% less likely to drop out (AOR = 0.58, 95% CI: 0.40–0.85).

Households with a family size of more than three had 1.96 times more likely to drop out of MCV2 than those with family size of less than three. (AOR = 1.96, 95% CI: 1.29–2.98). Finally Care givers who reported waiting time greater than 30 minutes at vaccination services were 3.34 time more likely to drop out MCV2 (AOR = 3.34, 95% CI: 1.86–5.90).

## Discussion

This study investigated the determinants of measles second dose (MCV2) dropout among children 24-35 months of age in Addis Ababa. The study shows that the odds of dropout is higher among those who didn’t receive reminder from health service delivery. This finding is in contrast with findings of community-based studies conducted in Ejere Woreda, Oromia Region, where lack of reminder significantly increased MCV2 dropout (19). Furthermore the finding is also in parallel with the study done in Kenya and Nigeria (27,28). An explanation for this might be due to reminder system such as Phone call, text message or home visits significantly improve immunization completion rate and the absence of it may contribute to forgetfulness or co or other competing priorities in urban settings where care giver often balance multiple responsibilities.

Lack of Counseling is also significantly associated with measles second dose dropout. As those who didn’t receive Counseling is more likely to drop out of measles second dose vaccination dropout. Findings from East Bale Zone, Tanzania, and Benin are in support of this, Which reported that lack of counseling from health care contributes to dropout of immunization schedule (21,29) The possible explanation for this is Counseling improves caregivers understanding of immunization schedule, alleviate misconceptions and enhances trust in vaccination programs.

Adverse event following immunization (AEFI) experience is also found to be a significant determinant of MCV2 dropout. The odds of drop out among Children with a prior adverse event following immunization had twice higher than those without prior adverse event following immunization. Similar findings have reported in Ethiopia and Nigeria, where AEFI deterred from finishing the vaccination series by fear of adverse events following vaccines (30,31). This might be due to post vaccination adverse event may be perceived as serious among caregivers with limited understanding which can create fear and anxiety, leading to avoid subsequent vaccination.

Mothers who have attended postnatal care visits were significantly associated with decrease drop out status. This protective effect align with evidence from East Bale Zone and the Ethiopian Demographic and Health Survey (20,21). Furthermore this finding is in consistent with study conducted in Wonago district of south Ethiopia, identified mothers who did not use PNC service after delivery of the child under study were 2 times more likely to have dropout children than mothers who did use PNC services (32). This Could be due to PNC visits provide an opportunity for health staff to counsel mothers, remind them, and emphasize the importance of good timing in vaccination, pointing out the advantage of integrating immunization promotion into maternal and child health care(33).

This study also shows Family size had a significance association with MCV2 dropout. The odds MCV2 dropout among Family of more than three were 1.96 times higher than those family size of less than three. The finding is in consistent with studies conducted in China and Ethiopia have also suggested that family size is greater with increased odds of vaccination dropout (14,34). The possible explanation for this might be large family size may limit caregivers available time, financial resources, and attention for follow up visit, Comparable findings were also reported in Nigeria and Nepal, where increase family burden was linked to vaccination dropout.(35–37). Addressing such socioeconomic barriers through community bases activities and flexible service hours could improve vaccination among large families. This may be attributed to competing caregiver time demands, scarce household resources, or difficulty in following up with multiple health visits.

Additionally, longer waiting times were found to be strongly associated with MCV2 drop out which the study shows that the odds of dropout among caregivers waiting more than 30 minutes for vaccination service is 3.34 times higher than those waiting for less than 30 minutes, there was a similar finding in Kenya, Benin, Tanzania and East Bale Zone where a long delay and long queues discouraged caregivers from following up for the subsequent doses (21,29,38,39). This might be due to mother/caregivers had experience of prolonged waiting time lead to dissatisfy and not being motive to get the second dose of vaccine for their children.

## Conclusion

From this study we can conclude that the factors affecting measles second dose drop out among children aged between 24-35 months In Addis Ababa are lack of reminder, poor counseling from health care workers, previous history of adverse event following immunization, family size of greater Than three and more than 30-minute waiting times at health facility for immunization service. Children whose caregivers did not receive reminder about vaccination, lack of counselling from health workers and adverse event following immunization (AEFI) had higher odds of dropout. Family size and waiting times at the health facility were significantly associated with increase odds of measles second dose (MCV2) vaccination dropout. On the other hand, post-natal care visit is associated with decrease odds of dropout.

## Limitation of the study

This study measured drop out status by using Case-control study design, as a result it’s susceptible to recall bias and social desirability bias.

## Declarations

Human ethics and consent to participate

This study is involving human participants was conducted in accordance with the ethical principles outlined in the Declaration of Helsinki.

Ethical approval was obtained from the Research and Ethics Board (REB) of Saint Paul hospital millennium medical college with reference number PM23/666 and protocol number of SPHMMC-ERC-092/25. Then it was submitted to Addis Ababa regional health bureau and clearance was obtained. During the data collection, informed consent was obtained from each respondent after explaining the objectives of the study and the rights of the respondent to participate or not in the study.

## Consent for publication

I Confirm I give my full consent that I have read and approved the final version of the manuscript and take full responsibility for its content. I affirm that I have the necessary authority to grant this consent and have obtained any required permissions for the publication of this work. Furthermore, I acknowledge that once published, the manuscript will be publicly accessible in print, online, and other formats as determined by the publisher. I confirm that the manuscript does not infringe upon any copyright, confidentiality, or legal obligations. Additionally, I agree that the publisher may make minor editorial modifications, provided they do not alter the integrity of the content.

## Availability of data and material

All the data generated in this study are included in this manuscript. The datasets used and analyzed to produce the current manuscript will be obtained from the corresponding author upon request.

## Authors’ contributions

BG designed the study, develop proposal and data collection tool, conducted the data analyses, write summary report and drafted the manuscript.

SH advise on proposal development, data collection, analysis, writeup, manuscript development and supervise overall process.

DT support collection of data and provide encouragement throughout study period. ZA advise on proposal development, data collection, analysis, writeup, manuscript development and supervise overall process.

## Data Availability

The datasets used and analyzed to produce the current manuscript will be obtained from the corresponding author upon request.

## Acknowledgement

First and for most I am sincerely grateful to Saint Paul’s Hospital Millennium Medical College School of Public Health, Department of Epidemiology for granting the opportunity to conduct this study. I would also like to extend my heartfelt appreciation to my advisors Mr. Simegnew Handebo and Dr. Zewdie Aderawt PhD, for their Constructive comments and continuous encouragement from the development of the research topic to the completion of this thesis. My sincere gratitude is also extended to the supervisors and data collectors who devoted their time, effort and professionalism during the data collection period. I would also like to express my profound appreciation to the study participants who generously shared their time to respond to the questions.

## Legends of table

Table 1: Sample size Calculation using a determinant variable with parameters by Epi info version 7.2.6.0 software. 2025

Table 2: Socio demographic characteristics of participants for determinants of measles MCV2 vaccination dropout among children aged 24-35 months of age, Addis Ababa Ethiopia 2025.

Table 3: Health Care system related factors for determinants of measles MCV2 vaccination dropout among children aged 24-35 months of age, Addis Ababa Ethiopia 2025.

Table 4: Child related factors for determinants of measles MCV2 vaccination dropout among children aged 24-35 months of age, Addis Ababa Ethiopia 2025.

Table 5: Mother / Care giver related determinants of measles MCV2 vaccination dropout among children aged 24-35 months of age, Addis Ababa Ethiopia 2025.

Table 6: Bivariable and multivariable analysis of measles MCV2 vaccination dropout among children aged 24-35 months of age, Addis Ababa Ethiopia 2025.

